# Telomere Biology Disorder Predisposition in Bone Marrow Failure and Clonal Hematopoiesis

**DOI:** 10.1101/2025.07.08.25331123

**Authors:** Daijing Nie, Xvxin Li

## Abstract

Telomere biology disorders (TBDs), exemplified by dyskeratosis congenita (DC), are characterized by genetic defects in telomere maintenance genes, leading to telomere attrition and multi-organ manifestations including bone marrow failure (BMF) and increased malignancy risk. This study aimed to evaluate the prevalence and clinical impact of rare possibly significant variations (PSVs) in telomere-related genes among patients with BMF and clonal hematopoiesis disorders. We analyzed 1320 patients diagnosed with aplastic anemia, myelodysplastic syndrome, acute myeloid leukemia, or acute lymphoblastic leukemia and identified 113 PSVs in 103 patients, significantly exceeding the prevalence in the general population (gnomAD). The most frequently mutated genes were *RTEL1* (37%) and *CTC1* (30%). While missense variants predominated, novel variants accounted for approximately 27.4%. Patients harboring PSVs exhibited significantly shorter telomeres compared to unaffected relatives, reinforcing telomere length as a critical functional biomarker. Telomere-mediated genetic anticipation was clearly evident: younger patients had notably shorter telomeres compared to their older first-degree relatives, reflecting cumulative generational telomere attrition and progressively severe phenotypes. Despite variability in clinical presentations—with many patients lacking classical mucocutaneous or fibrotic manifestations—telomere shortening provided robust onset information. Our findings emphasize the importance of integrating genetic testing and telomere length measurement into clinical practice for early diagnosis, personalized risk stratification, and tailored management, including reduced-intensity conditioning transplantation and emerging targeted therapies.

## Introduction

Telomeres, consisting of repetitive (TTAGGG)_*n*_ sequences bound by the shelterin complex, protect chromosome ends from inappropriate DNA-damage responses[1, 11]. Due to incomplete replication, telomeres shorten progressively during each cell division[2]. Critically short telomeres trigger ATM-mediated DNA damage signaling, activating p53 or p16/RB pathways to induce senescence or apoptosis, thus preventing malignant transformation[9, 3]. Conversely, dysfunctional telomeres can cause chromosomal end-to-end fusion, genomic instability, and ultimately, malignant evolution[1, 7].

Germline loss-of-function variants in telomere maintenance genes such as *TERT, TERC, DKC1, RTEL1*, and others result in telomere biology disorders (TBDs), typified by dyskeratosis congenita (DC)[10]. Classical DC is characterized by childhood-onset mucocutaneous manifestations, bone marrow failure (BMF), and increased cancer susceptibility. However, many patients exhibit cryptic adult-onset phenotypes, including idiopathic pulmonary fibrosis (IPF), liver fibrosis, isolated cytopenias, or malignancies, often lacking classical features. Telomere-mediated genetic anticipation—progressively shorter telomeres leading to earlier and more severe disease in successive generations—is a hallmark of these disorders[10].

Telomere length assessment complements genetic testing by providing crucial functional and prognostic information, guiding clinical management and genetic counseling[7, 12]. Current treatments for TBDs emphasize supportive care, careful use of androgens, reduced-intensity conditioning hematopoietic stem cell transplantation (HSCT), and potential novel strategies such as telomerase activation or gene therapy[12, 8, 13, 5]. However, key questions remain, including the prognostic significance of specific gene variants and telomere length dynamics during disease progression.

This study integrates genetic profiling, telomere length assessment, and clinical data to clarify the clinical implications of telomere-related gene variants, aiming to refine diagnostic criteria, enhance patient risk stratification, and inform targeted therapeutic strategies for telomere-associated bone marrow failure and hematologic malignancies.

## Materials and Methods

### Patient Cohort

We retrospectively reviewed the medical records of patients who were hospitalized between April 2015 and December 2020. Diagnostic categories were established as follows:

- **AA** was defined by the presence of cytopenia affecting at least two peripheralblood lineages, reticulocytopenia, and hypocellular bone marrow confirmed by Wright–Giemsa smears and/or haematoxylin–eosin histopathology.
- **AML, MDS and ALL** were classified according to the 5th edition of WHO criteria for tumours of haematopoietic and lymphoid tissues[4].

The electronic medical record was reviewed in detail for (i) disease course, (ii) laboratory and histopathological data and (iii) personal and familial features suggestive of an inherited bone-marrow failure or telomere biology disorder. All procedures were approved by the Hebei Yanda Lu Daopei Hospital Ethics Committee and conformed to the principles of the Declaration of Helsinki (1975, revised 2013). Written informed consent was obtained from every participant prior to enrolment.

## Genetic Variant Identification

Amplicon-based high-throughput sequencing was performed as previously described [6]. Variant calling was carried out using Torrent Variant Caller (TVC v5.0-13, Thermo Fisher Scientific), and high-confidence variants were annotated with ANNOVAR. Variants with a minor allele frequency (MAF) *≥* 0.1% in the general population were excluded based on the Exome Aggregation Consortium (ExAC) and Genome Aggregation Database (gnomAD).

The pathogenicity of germline missense variants was evaluated using six in silico prediction tools: SIFT, PolyPhen-2, PROVEAN, FATHMM, MutationTaster, and MutationAssessor. To account for potentially hypomorphic alleles and incompletely penetrant variants, a relaxed criterion was applied: any variant predicted as deleterious or possibly deleterious by *≥* 3 out of 6 algorithms was classified as a rare potentially significant variant (PSV) and included in further analysis.

For splice site variants, four splicing prediction tools were employed: GeneSplicer, Human Splicing Finder, NetGene2, and FSPLICE. A variant was considered potentially significant if *≥* 2 out of 4 tools predicted a likely effect on splicing.

All reported variants were confirmed as germline through Sanger sequencing of fingernail DNA and/or pedigree analysis.

## qPCR Telomere Length Assay

Telomere length was measured using a quantitative PCR (qPCR)-based assay. Cryopreserved DNA extracted from peripheral blood mononuclear cells (PBMCs) of patients and their first-degree relatives was used for analysis. PCR amplification was performed separately for telomeric repeats (*TEL*) and a singlecopy reference gene (*SCR*, e.g., 36B4). The relative telomere length (T/S ratio) was calculated using the quantification cycle (*Cq*), defined as the PCR cycle at which the amplification fluorescence signal exceeds a defined threshold, according to the formula:

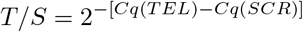

An age-dependent reference standard was generated from PBMC DNA samples collected from 1,656 randomly selected healthy individuals aged between 1.5 and 98 years. Age-specific percentile curves (1st, 10th, 50th, 90th, and 99th percentiles) were derived by fitting telomere length data to a one-phase exponential decay model. All qPCR analyses included appropriate internal standards and quality controls to ensure reproducibility.

## Statistical Analysis

Variant frequencies between groups were compared using Fisher’s exact twotailed test. The cumulative incidence method was employed to analyze disease development. Statistical analyses were performed using Python 3.12.9, and a two-sided *p*-value less than 0.05 was considered statistically significant.

## Results

### Demographics and Clinical Features

A total of 1320 patients diagnosed with AA or AA-PNH, MDS, AML, or ALL were included, each with comprehensive medical records. The median age at onset, age range, sex ratio, and the case numbers of disease subgroups are detailed in Table 1. Except for 16 individuals of West Asian and 3 individuals ofm Middle Eastern ancestry, all other patients were of East Asian ancestry.

**Table 1:** Demographic and Clinical Characteristics of the Patient Cohort.

| Group | Median age at onset, year (range) | Sex ratio (male/female) | Case number |
| --- | --- | --- | --- |
| Pediatric | 5 (0–14) | 461/340 | 801 |
| Adolescent and young adult | 24 (15–39) | 215/163 | 378 |
| Adult | 48 (40–65) | 69/72 | 141 |
| AA/AA-PNH | 9 (0–63) | 220/171 | 388 |
| MDS | 29 (1–66) | 69/64 | 133 |
| AML | 13 (0.5–65) | 197/163 | 360 |
| ALL | 7 (0–64) | 250/175 | 425 |
| Others | 5.5 (1–19) | 9/5 | 14 |
| Total | 11 (0–65) | 745/575 | 1320 |

Others include Fanconi anemia, lymphoma, and mixed phenotype leukemia.

### Features of Variants

In total, 113 variants meeting the PSV criteria were identified in 103 patients (113/1320, 8.56%; Table S1, Table S2). The distribution of variants by gene and mutation type is illustrated in Figure 1. The most frequently mutated gene was *RTEL1* (37%), followed by *CTC1* (30%). Due to the single-copy nature of the *DKC1* gene in the human genome, its variants constituted the smallest proportion (1%). Consistent with other inherited disorders, missense mutations were most common (85%), followed by frameshift mutations leading to stop-gain (7%). Nonsense mutations and small insertions/deletions (InDels) accounted for 3% each, and telomerase RNA variants, unique to this class of disorders, represented 2% of the total[6]. Among these 113 PSVs, 31 were classified as novel variants, not previously reported in population databases or clinical variant repositories. Notably, the *RTEL1* c.1956C*>*A/p.S652R variant was identified in two unrelated patients of East Asian ancestry, representing a recurrent variant. All employed in silico prediction algorithms consistently indicated this variant as pathogenic or highly deleterious, suggesting it may represent an East Asian-specific pathogenic variant (Table S2).

**Figure 1:**
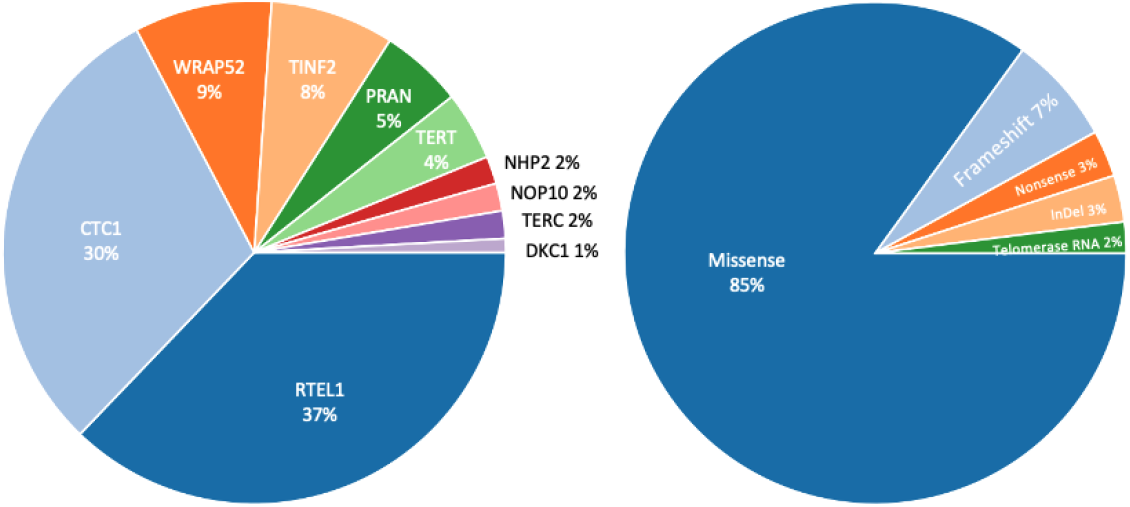
Distribution of identified variants by gene (left) and mutation type (right).

We then compared PSVs between our patient cohort and the general population data from the Genome Aggregation Database (gnomAD), which includes genomic data from 807,162 individuals. Using identical criteria, the overall PSV carrier rate for the 10 genes studied was 0.96% in the gnomAD cohort. Compared to the general population, our patient cohort as a whole, and each disease subgroup individually, exhibited significantly higher PSV carrier rates. These results underscore the significant contribution of telomere-related gene variants to bone marrow failure and clonal hematopoiesis disorders (Table 2).

**Table 2:** Comparison of PAV carrier rate between patient cohorts and gnomAD population.

| Group | Total | PSV | PSV% | OR | 95%CI | p |
| --- | --- | --- | --- | --- | --- | --- |
| AA/AA-PNH | 388 | 41 | 10.57 | 11.201 | 8.012–15.611 | $5.99 \times 10^{-26}$ |
| MDS | 133 | 10 | 7.52 | 5.731 | 2.732–11.928 | $3.27 \times 10^{-4}$ |
| AML | 360 | 28 | 7.78 | 7.699 | 5.136–11.505 | $3.73 \times 10^{-14}$ |
| ALL | 425 | 34 | 8.0 | 8.117 | 5.635–11.660 | $9.01 \times 10^{-18}$ |
| Total | 1306 | 113 | 86.52 | 8.597 | 7.434–9.937 | 0 |

Patient DC 016, diagnosed with aplastic anemia (AA), carried three variants in the *TERT* gene. Variant *TERT* c.3346G*>*C was inherited from the mother, while variants *TERT* c.2839T*>*C and c.1796G*>*A were inherited from the father. Patient DC 041, also diagnosed with AA, carried three variants: a *TINF2* c.1285C*>*G variant, an *RTEL1* c.1721G*>*T variant inherited from the mother, and an *RTEL1* c.3073 3074insT/p.A1025Vfs*66 variant inherited from the father. Both patients exhibited the classical dyskeratosis congenita triad of skin pigmentation, oral leukoplakia, and nail dystrophy. Additionally, patient DC 002 carried a heterozygous *TINF2* c.844C*>*T/p.R282C variant, and patient DC 010 carried a heterozygous *TINF2* c.849delC/p.T284Qfs*33 variant. These two patients also presented clinically with the classical triad and bone marrow failure. The remaining patients lacked characteristic clinical manifestations and radiographic evidence of pulmonary fibrosis.

## Telomere Length

We analyzed telomere lengths in 229 patients harboring variants in telomere-associated genes, along with 143 of their first-degree relatives. The median age of the patients was 13 years, ranging from 1 to 64 years. The median age of the relatives was 35 years, ranging from 10 to 68 years. These variants in these patients were not necessarily classified as PSV according to our study criteria, but we performed comprehensive telomere length assessments in all 103 confirmed PSV carriers. Despite the majority of these 229 patients lacking typical clinical manifestations (such as the classical triad or organ fibrosis), significant telomere attrition was identified. Of these patients, 30/229 had telomere lengths below the 1st percentile of age-matched healthy references, including 14/30 who were confirmed PAV carriers. Additionally, 83/229 patients had telomere lengths below the 10th percentile, among whom 31 carried confirmed PSVs. Overall, among 103 patients carrying PSVs, 45 exhibited shortened telomeres.

In contrast, among 143 first-degree relatives, only one individual (1/143) had telomere length below the 1st percentile; notably, this individual carried a heterozygous *TERT* c.2839T*>*C/p.S947P variant. Additionally, 10/143 relatives had telomere lengths below the 10th percentile, and 5 of these individuals were identified as PAV carriers (Figure 2; Table S2).

**Figure 2:**
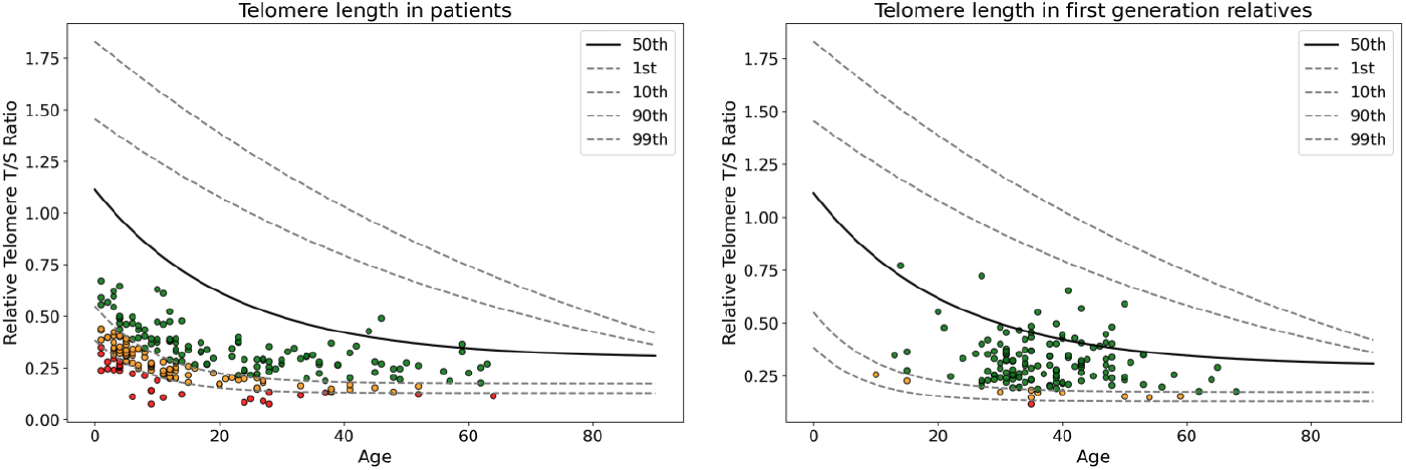
Relative telomere T/S ratio in of patients (left) and first degree relatives (right). Red dots represent the ones which are shorter than the 1st percentile of normal population; orange dots represent the ones which are shorter than the 10th percentile of normal population; and green dots represent the ones with normal length which are longer than the 10th percentile of the reference.

## Cumulative Incidence

We evaluated the impact of germline pathogenic or likely pathogenic variants (PSVs) in telomere-related genes on disease onset across patient subgroups (Figure 3). In the aplastic anemia group, PSV carriers exhibited significantly earlier disease onset compared to non-carriers (median age: 8 vs. 12 years, *p* = 0.018), indicating a substantial contribution of germline variants to early-onset marrow failure. In contrast, PSV status did not significantly influence the age of onset for patients with myelodysplastic syndrome (MDS, *p* = 0.56), acute myeloid leukemia (AML, *p* = 0.89), or acute lymphoblastic leukemia (ALL, *p* = 0.63). These findings suggest that while telomere-related PSVs markedly predispose individuals to early marrow failure, malignant transformation in MDS and acute leukemias involves more complex mechanisms, likely influenced by additional germline variants or somatic mutations.

**Figure 3:**
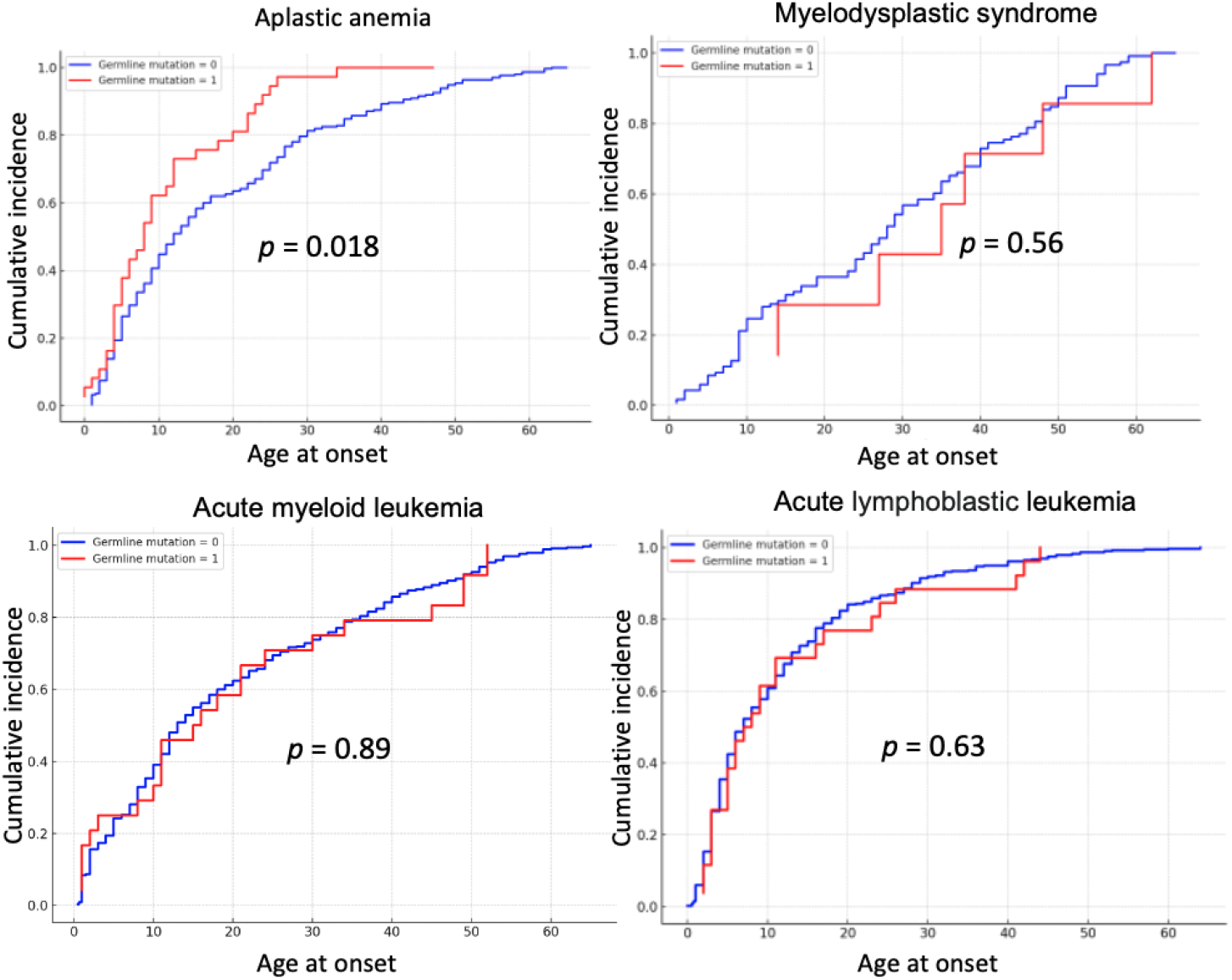
Cumulative incidence curves comparing disease onset in patients with (red lines) and without (blue lines) germline telomere-related variants across aplastic anemia, myelodysplastic syndrome, acute myeloid leukemia, and acute lymphoblastic leukemia subgroups.

## Discussion

In this study, we observed a significant enrichment of PSVs in telomere-related genes among patients with bone marrow failure and malignant clonal hematopoiesis, underscoring their critical contribution to disease onset and progression. Despite limitations of in silico prediction methods, the strong correlation observed between variant status, clinical phenotype, and telomere shortening supports our PSV criteria’s clinical relevance. Telomere length measurements provided critical functional insight that genetic sequencing alone could not fully capture. Even in patients without classic dyskeratosis congenita-associated clinical features such as mucocutaneous triad or overt organ fibrosis, severe telomere attrition was frequently observed. Our findings reinforce the diagnostic and prognostic utility of integrating genetic variant analysis with telomere length assessment in clinical practice. Such integration allows early identification of at-risk individuals, accurate diagnosis in cryptic or atypical cases, and personalized management strategies.

Patients PSVs demonstrated a notably higher cumulative incidence of marrow failure at younger ages compared to non-carriers, underscoring a critical etiological contribution of telomere dysfunction. Specifically, in the aplastic anemia subgroup, PSV carriers had a substantially earlier disease onset (median age 8 versus 12 years, *p* = 0.018), consistent with the phenomenon of genetic anticipation, whereby successive generations exhibit progressively shorter telomeres, earlier disease manifestation, and more severe phenotypes. This cumulative generational attrition of telomeres is a hallmark of telomere biology disorders and emphasizes the need for early genetic counseling and risk assessment within affected families.

Interestingly, our cumulative incidence analysis revealed no significant association between telomere-related PSVs and onset age in patients with myelodysplastic syndrome (MDS), acute myeloid leukemia (AML), or acute lymphoblastic leukemia (ALL). This lack of correlation suggests that malignant transformation in these contexts is likely driven by complex interactions involving additional germline predispositions and acquired somatic mutations, rather than telomere dysfunction alone. Recent genomic studies support this notion, demonstrating that the development of hematologic malignancies typically requires cooperating mutations in critical signaling pathways or epigenetic regulators beyond the initial telomere-related genetic insults.

In conclusion, comprehensive evaluation incorporating germline variant profiling, telomere length assessment, and clinical phenotype characterization is essential for optimizing diagnostic accuracy, informing precision medicine approaches, and ultimately improving clinical outcomes for patients with telomere biology disorders.

## Data Availability

All data produced in the present study are available upon reasonable request to the authors

